# Characteristics of People with Type I or Type II Diabetes with and without a History of Homelessness: A Population-based Cohort Study

**DOI:** 10.1101/2022.08.11.22278127

**Authors:** Kathryn Wiens, Li Bai, Peter C Austin, Paul E Ronksley, Stephen W Hwang, Eldon Spackman, Gillian L Booth, David JT Campbell

## Abstract

**Introduction:** Homelessness poses unique barriers to diabetes management. Population-level data on the risks of diabetes outcomes among people experiencing homelessness are needed to inform resource investment. The aim of this study was to create a population cohort of people with diabetes with a history of homelessness to understand their unique demographic and clinical characteristics and improve long-term health outcomes.

**Methods:** Ontario residents with diabetes were identified in administrative hospital databases between 2006 and 2020. A history of homelessness was identified using a validated algorithm. Demographic and clinical characteristics were compared between people with and without a history of homelessness. Propensity score matching was used to create a cohort of people with diabetes experiencing homelessness matched to comparable non-homeless controls.

**Results:** Of the 1,455,567 patients with diabetes who used hospital services, 0.7% (n=8,599) had a history of homelessness. Patients with a history of homelessness were younger (mean: 54 vs 66 years), more likely to be male (66% vs 51%) and more likely to live in a large urban centre (25% vs 7%). Notably, they were also more likely to be diagnosed with mental illness (49% vs 2%) and be admitted to a designated inpatient mental health bed (37% versus 1%). A suitable match was found for 5219 (75%) people with documented homelessness. The derived matched cohort was balanced on important demographic and clinical characteristics.

**Conclusion:** People with diabetes experiencing homelessness have unique characteristics that may require additional supports. Population-level comparisons can inform the delivery of tailored diabetes care and self-management resources.

## Background

Homelessness is a pressing health concern that has important implications for diabetes care. The prevalence of diabetes is similar for homeless and non-homeless groups (1, 2); however, living without a home poses unique barriers to self-management. Common barriers include a lack of access to treatment and testing supplies, scarcity of nutritious foods, and difficulty storing medications (3, 4). Many people experiencing homelessness have competing priorities, such as procuring food, shelter, or income, that are often placed before their medical needs. They also report mistrust of the medical system, which further impedes access to care (5). Over time, these barriers can contribute to hyper- and hypoglycemia (4, 6). One study reported suboptimal glycemia in 44% of patients experiencing homelessness at community shelter clinics, with nearly one-third having glycated hemoglobin values above 9% (7). Hyperglycemia is an important risk factor for complications, such as cardiovascular disease and mortality (8, 9).

Diabetes complications are more common among people facing social disadvantage, such as housing instability and homelessness. Unstable housing was associated with 5 times higher odds of a self-reported hospital visit in the past year compared to stable housing, based on data from federally funded safety-net clinics in the United States (10). In Canada, lower socioeconomic status was also linked with higher rates of hospital service use for glycemic emergencies (11). Other complications, such as retinopathy and heart disease, were higher in groups with more social disadvantage in the United Kingdom (12), and a small study of people with diabetes in France reported 15 times higher amputation rates for people experiencing homelessness than the general population (13). Inpatient care for diabetes complications is expensive (14) and can possibly be avoided with adequate access to treatment and self-management resources (15).

To date, much of our knowledge of diabetes outcomes among people experiencing homelessness is from qualitative studies. Our ability to study health outcomes at the population level has improved with advancements in administrative data linkage; however, the application of these data to study homelessness has been limited. One study using State Inpatient Databases reported that youth with diabetes experiencing homelessness have higher readmission rates and length of hospital stay, and lower rates of diabetic ketoacidosis and hospital costs than non-homeless youth with diabetes in New York (16). Another study using Veterans Health Administration data showed higher odds of elevated glycated hemoglobin levels (>8%) for Veterans with diabetes who are homeless compared to housed (17). There is a need for additional population-level data on long-term disparities in health outcomes for people with diabetes experiencing homelessness. This information would help to inform planning for and delivery of tailored services that improve access to diabetes care and self-management resources.

In Ontario, all residents are covered for medically necessary services under the Ontario Health Insurance Plan (OHIP). Information on their use of health care services is available in centralized provincial healthcare databases. Previous researchers have created a validated registry of all Ontario residents with diabetes, based on a history of hospital admissions or outpatient physician visits for diabetes (18). In 2019, an algorithm was validated to identify people experiencing homelessness using Ontario healthcare databases (19). The availability of these registries and databases creates an opportunity to identify a large population-based cohort of people with diabetes with a documented history of homelessness.

Given the current knowledge gaps and recent advances in identification of diabetes and homelessness at a population-level, the first objective of this study was to create a population cohort of Ontario residents with diabetes who have a history of homelessness to understand how their demographic and clinical characteristics differ from people without a history of homelessness. By highlighting these differences, this cohort provides an opportunity to tailor diabetes care to meet the unique needs of people experiencing homelessness. The second objective was to assess whether propensity score methodology could be used to develop a matched cohort of people with a history of homelessness and non-homeless controls to enable population-level comparisons of long-term diabetes outcomes by homeless status.

## Methods

### Data sources

In Ontario, Canada, residents are eligible for healthcare coverage under OHIP, a single-payer insurance system paid by the Ontario Ministry of Health and Ministry of Long-Term Care. Coverage is provided for all medically necessary healthcare services, including hospital and physician services, laboratory tests, and medications received in hospital. Additional medication coverage is available for residents over 65 years old and people qualifying for social assistance benefits. Administrative data from these services are routinely collected and stored at ICES, a not-for-profit research institute, for the purpose of health system analysis, evaluation, and decision support. The Registered Persons Database is a registry of Ontario residents covered by OHIP. Each resident is assigned a unique ICES key number to enable linkage across ICES databases.

We used these administrative databases to identify a cohort of adults (18 years or older) living with diabetes who use hospital services in Ontario. Each eligible person required a valid health card number (OHIP eligibility) and a diagnosis of diabetes. The Ontario Diabetes Dataset (ODD) is a registry of all Ontario residents who had at least one hospital admission for diabetes *or* two outpatient physician claims for diabetes within two years (18). Hospital databases have the most extensive list of homeless-identifying data elements in Ontario (19), so each person with diabetes required at least one inpatient admission or emergency department visit during the accrual period (April 1, 2006, to March 31, 2019). Supplemental Table S1 describes the data used to create the cohort. These datasets were linked using unique encoded identifiers and analyzed at ICES.

### Exposure: homeless status

Homeless status was defined for the cohort of adults living with diabetes in Ontario based on documentation in hospital records. Supplemental Table S2 describes the databases and indicator codes used to identify homelessness. In 2019, a set of algorithms were validated to identify homelessness in ICES administrative data using an external cohort of adults experiencing homelessness (19, 20). The high positive likelihood ratios and specificity of these algorithms provide confidence that the people who are identified as homeless in administrative data are truly homeless. The low sensitivity (18-35%) recognizes that many people experiencing homelessness will not be identified with the existing administrative data algorithms (19).

People were classified as having a history of homelessness if they had at least one hospital encounter with documented homelessness during the accrual period (Supplemental Figure S1). Index dates were assigned separately for people with and without a history of homelessness. For each person identified as homeless, the index date was the date of discharge from their first hospital encounter with documented homelessness. Since the first encounter with documented homelessness could occur at any time during the follow-up period, we forced the non-homeless controls to have a similar distribution of index dates. For each non-homeless control, we assigned an index date as the date of discharge from a hospital encounter that was randomly selected according to the distribution of index dates in the group with documented homelessness.

#### Variables of interest

The variables on which homeless and non-homeless groups were matched include demographic and clinical characteristics that are associated with homelessness and risk factors for diabetes outcomes, as shown in Supplemental Figure S1.

Index year, age, type of hospital encounter, geographical region, and presence of mental illness or substance use disorders were included as hard match variables to ensure balance was achieved between groups. Age was a continuous variable ascertained from the Registered Persons Database. Type of hospital encounter was a set of indicator variables for at least one hospital visit to a designated inpatient mental health bed (OMHRS), other inpatient admission (DAD), or emergency department visit (NACRS) within 6 months of index date. Region of residence was captured using nine Local Health Integration Networks (LHINs) as a measure of geographical location. Presence of a mental illness or substance use was an indicator variable for any hospital encounter for mental illness or addictions in the year before index date.

The variables included in the propensity score for homelessness were sex, duration of diabetes, type 1 diabetes, and past hospitalization for acute myocardial infarction, heart failure, or stroke prior to index date. Sex was ascertained from the Registered Persons Database. Duration of diabetes was calculated using date of entry into the ODD (< 2 years, 2-5 years, 5-10 years, and ≥10 years). Type of diabetes was classified using an algorithm at ICES, which relies on age of diagnosis and prescription of insulin or insulin pump therapy without any oral antihyperglycemic agents to identify type I diabetes (21). Past myocardial infarction, stroke, or heart failure were identified using ICD-10 codes as listed in DAD. These conditions were included to ensure the baseline risk for major adverse cardiovascular events was comparable between groups.

Additional categories of mental illness and chronic conditions were used to describe the cohort but were not included in the match criteria. Type of mental illness and substance use were assessed as a set of indicator variables for psychotic disorder, bipolar disorder, other mental disorders, alcohol use, and drug use. Other chronic conditions were chronic obstructive pulmonary disease, rheumatic disease, dementia, hemiplegia or paraplegia, HIV, liver disease, peptic ulcer disease, cancer, and renal disease.

#### Statistical Analysis

Demographic and clinical characteristics were compared between people with and without a history of homelessness using standardized differences and p-values from Kruskal-Wallis tests (median) for continuous variables and Chi-Squared tests for categorical variables.

A propensity score for homelessness was estimated using a logistic regression model regressed on sex, duration of diabetes, type 1 diabetes, and past major adverse cardiovascular events, as defined above. Each patient with documented homelessness was hard matched to a non-homeless control on index year, age within 5 years, a set of indicator variables for type of hospital encounter, geographical region, presence of mental illness or substance use disorder, and the logit of the propensity score within 0.2 calipers using greedy matching without replacement (22). The suitability of the match was examined using standardized differences and variance ratios (23). A standardized difference of greater than 0.1 indicates covariate imbalance. For continuous variables, the variance ratio is expected to be close to 1 if balance was achieved between groups.

A secondary analysis was undertaken to determine if any excess risk of adverse outcomes was due to income disparities rather than homelessness specifically. In Ontario, residents are allocated to a neighborhood income quintile based on their six-digit postal code as reported in administrative records. Each person identified as homeless during the accrual period was exclusively matched to a non-homeless control from a low-income neighbourhood.

All analyses were completed using SAS software.

## Results

### Cohort description

Figure 1 describes the study inclusion criteria. Initially, there were 2,324,716 people with diabetes identified in the ODD up until March 31, 2019. Of these, 1,455,567 (63%) had at least one hospital encounter after April 1, 2006. People less than 18 years old on March 31, 2019 (n=43,751) or who were living outside Ontario for the duration of the study (n=191) were excluded. The remaining cohort consisted of 1,411,625 eligible adults with diabetes who used hospital services in Ontario between April 1, 2006, to March 31, 2019.

**Figure 1:**
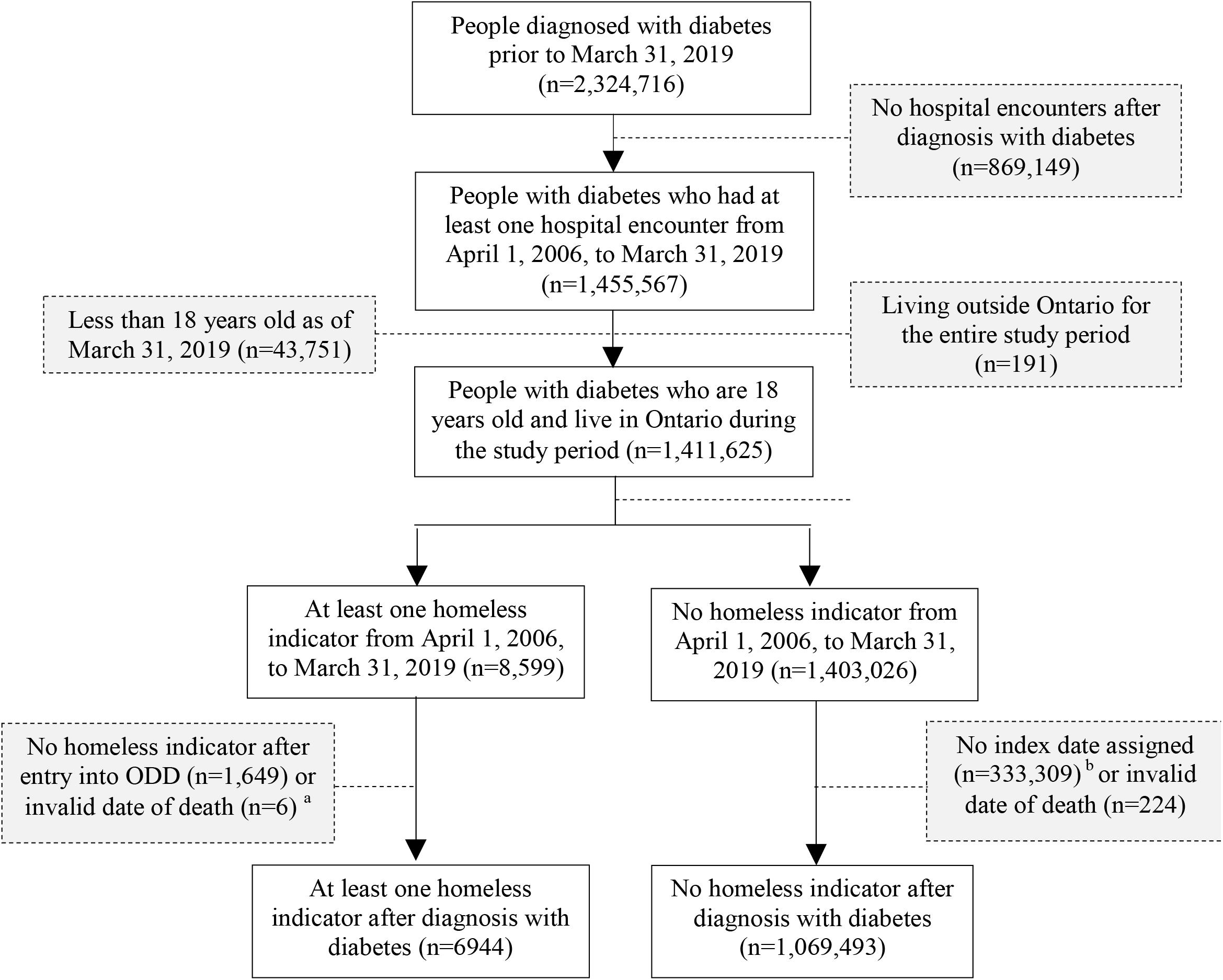
Inclusion criteria for people with diabetes who use hospital services a. There were <6 invalid deaths (index date after date of death). Exclusion counts were modified to supress small cells. b. An index date could not be assigned for these individuals because they did not have a hospital encounter that matched the distribution of index dates among the people with a history of homelessness.

A history of homelessness was documented for 8,599 eligible people with diabetes during the study period, which is approximately 0.7% of the eligible cohort. Index dates were assigned to people with a history of homelessness as the date of their first hospital encounter with documented homelessness after their diabetes diagnosis. Notably, 1,649 people were excluded since their only hospitalizations happened prior to their diagnosis with diabetes. Therefore, index dates were only assigned to 6950 people in the homeless group. The percent of people with index dates assigned in the first 10 years was relatively consistent at 5-7% per year from 2006 to 2015. Approximately 8% of the cohort was assigned an index date in 2016, 9% in 2017, 16% in 2018, and 4% in 2019.

Index dates were assigned to 1,069,717 (76%) people without a history of homelessness for comparability with the distribution of index dates in the homeless group (Table 3). This led 331,626 people to be excluded. There were an additional 6 people in the homeless cohort and 224 people in the non-homeless cohort excluded due to invalid date of death – the assigned index date was after the persons documented date of death. The final sample was comprised of 6944 people with a history of homelessness and 1,069,493 people without a history of homelessness.

### Characteristics of people with diabetes, compared by homeless status

Table 3 highlights the differences in demographic and clinical characteristics for people with diabetes who have a history of homelessness compared to those who do not, prior to matching. People with a history of homelessness were younger, more likely to be male (66% versus 51%) and more likely to live in a large urban centre (i.e., Toronto Central; 25% versus 7%). They were also more likely to be diagnosed with mental illness, such as psychotic disorders (24% versus 1%), and be admitted to a designated inpatient mental health bed (37% versus 1%). In contrast, they were less likely to have previously had a diagnosed myocardial infarction (12% versus 15%) or heart failure (15% vs 16%), and just as likely to have had a stroke (10%).

### Propensity score match results

For the primary match of people with a history of homelessness to non-homeless controls, a suitable control was found for 5219 (75%) of the 6944 people with a history of homelessness (Figure 2). Standardized differences were less than 0.1, which indicate that covariate balance was achieved between groups (Table 4). The characteristics of the matched cohort largely reflect the characteristics of people with documented homelessness prior to the match. However, there were some differences due to the exclusion of 1725 people without a suitable match. The unmatched group of people with documented homelessness were younger (50 versus 55), more likely to be female (38% versus 32%), and less likely to live in Toronto Central (20% versus 27%) than the matched group of people with documented homelessness. They were also more likely to be admitted to a designated mental health bed (76% versus 24%) and have a psychotic (81% versus 19%), bipolar (21% versus 10%), or other mental disorder (58% versus 43%). The matched and unmatched groups are further compared in Supplemental Table S3.

**Figure 2:**
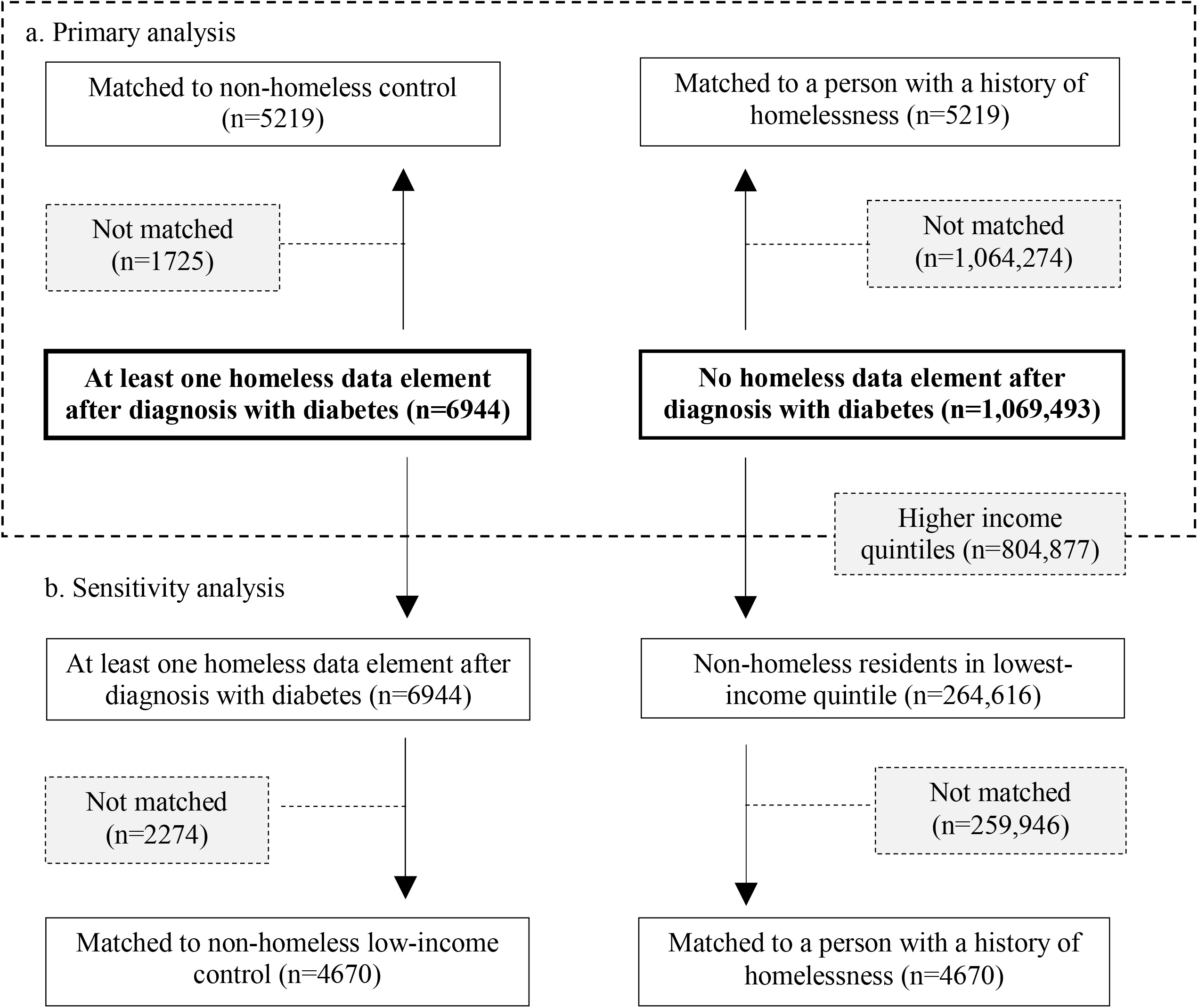
Flow diagram to depict the match procedure for people with a history of homelessness matched to non-homeless controls (primary) and non-homeless low-income controls (secondary)

For the secondary analysis comparing people with a history of homelessness to non-homeless low-income controls, 4670 matched pairs were found (67%). The distributions of demographic and clinical characteristics were similar for both matches (Table 4), except for the type of hospital encounter within 6 months of index date. Notably, the proportion of people admitted to a designated mental health bed was lower in the secondary match (19% versus 24%).

## Discussion

Diabetes management requires consistent care and behavioural modifications to achieve optimal glycemia. People experiencing homelessness have unique characteristics and healthcare needs, which may necessitate additional supports to manage their diabetes. Supportive housing initiatives must remain a priority; however, healthcare providers have an important role in the support of diabetes care and self-management for people experiencing homelessness.

Over the past few decades, improvements in data linkage have contributed to an increased use of administrative data to study population-level health outcomes; however, until recently, this has not been done for people experiencing homelessness. This study identifies a population of people with diabetes who use hospital services in Ontario, Canada, and describes the characteristics of a matched cohort of people with a history of homelessness and comparable non-homeless controls.

### Summary of findings

Approximately 0.7% of the cohort had a documented history of homelessness. This is higher than a previous estimate of 0.5% of the Ontario adult population with documented homelessness (19). Our study was restricted to people with diabetes who use hospital services, which likely contributed to this difference. In another study of Veterans with type 2 diabetes, 0.7% reported being homeless (17). Further, people experiencing homelessness are more likely to use hospital services than people who are housed (24, 25). Therefore, the prevalence of homelessness may appear higher within the subgroups of people who use hospital services.

Among the people in our cohort with documented homelessness, the largest proportion was identified in 2018 (16%). This was the year that the Canadian Institute for Health Information made it mandatory to use ICD-10 codes to document homelessness in emergency department and hospital data across Canada (26). We observed a slightly higher proportion of people identified as homeless in the two years leading up to this change as well. The number of people identified as homeless in Ontario administrative databases steadily increased over time (19, 27), which may represent changes in prevalence of homelessness or a shift in use of health services.

The demographic characteristics of the people with diabetes experiencing homelessness in our cohort were comparable with previous studies (20). Most people were in the 45-to-64-year age group (51%), with an average age of 54 years. Males contributed the largest proportion of people experiencing homelessness, as did people living in Toronto Central. Urban centres are common locations for people experiencing homelessness to access health services; however, there has been a growing number of people identified as homeless outside of Toronto in recent years (27).

People with a history of homelessness were more likely to access care for psychiatric conditions and addictions; however, the proportions observed in our study were higher than previous estimates of mental illness prevalence within the homeless population (20, 28). The requirement for people in our study to have a hospital encounter likely contributed to the high prevalence of mental illness for the people experiencing homelessness. Notably, one-third of people identified as homeless were admitted to a designated mental health bed within 6 months of their index date.

The people with documented homelessness were more likely to have a mental illness and less likely to have cardiovascular disease compared to non-homeless controls. In previous studies, the prevalence of cardiovascular disease is higher for people experiencing homelessness than non-homeless controls (29). This difference may reflect the younger age of our cohort. Cardiovascular mortality is also higher for people experiencing homelessness, with late presentation to care being a suggested reason for this disparity (30).

These findings reflect the major differences in the homeless and non-homeless populations and reinforce the need to match on demographic and clinical characteristics when studying diabetes outcomes. In this study, we successfully derived a matched cohort, achieving balance of the important characteristics that may confound the relationship between homelessness and diabetes complications. In future work, the covariates included in the propensity score match procedure may need to be modified for other diabetes outcomes, such as healthcare costs.

### Strengths

Administrative healthcare databases are a valuable source of longitudinal information that can be leveraged to study diabetes outcomes at the population level. The use of these databases is an important strength of this work, as we were able to capture a patient’s complete health data from their time of entry into the cohort until their date of death. This is particularly important when considering the challenges inherent in maintaining contact with a transient group of people over time. The use of centralized province-wide administrative databases mitigated the loss to follow up that occurs in prospective cohort studies with primary data collection from people living without a stable residence (31). It also enabled us to capture all provincial residents who used hospital services during our study period.

Another strength is the application of a recently validated homeless algorithm to identify people with documented homelessness in administrative databases. The high specificity and positive likelihood ratio of this algorithm increased our confidence that people identified as homeless during encounter were correctly classified (19). The algorithm sensitivity was low, which implies many people experiencing homelessness cannot be identified. However, we further restricted our cohort to people who used hospital services. Given nearly two-thirds of false negatives in the validation study were classified as housed by default due to no hospital encounter during the period of observation (19), we expect the sensitivity in our patient cohort was likely higher.

Finally, this study lays the foundation for future research to compare diabetes outcomes between people with and without a history of homelessness. For instance, the long-term impact of the COVID-19 pandemic on diabetes care for people experiencing homelessness has not yet been understood. The cohort described herein can be applied to study diabetes care and complications before, during, and after the pandemic.

### Limitations

There are some limitations to consider. First, it is likely that some people experiencing homelessness were misclassified as housed during the encounter. However, due to the low estimated prevalence of homelessness in the general population (0.5%) (29), the effect of some homeless individuals being coded as non-homeless is likely to be minimal (for example, given the 0.5% prevalence, we might expect that 26 of our 5219 non-homeless controls may in fact be homeless). Second, homelessness is transient; however, administrative healthcare data are not validated to classify people as homeless or housed in a time-dependent manner. Instead, people were defined as having a history of homelessness if they were identified as homeless during at least one encounter over the study period. Therefore, it is possible that some people identified as homeless were not homeless for the entire study, which may lead to more similarities in housing status between the homeless and non-homeless groups over time. However, it is expected that people with a history of homelessness will continue to experience long-term social and structural challenges after formally exiting homelessness. Third, since administrative data variables to identify homelessness are based on hospital encounter data, our study could not identify people experiencing homelessness who do not use hospital services. Therefore, caution must be taken when generalizing findings to people with diabetes who do not access hospital services.

It is important to understand the characteristics of people experiencing homelessness who could not be matched to a non-homeless control, as these people will not be captured in any analyses that use this cohort. Mental illness was an important factor that impacted our ability to find non-homeless matches for people experiencing homelessness. Identifying homelessness from psychiatric admissions in OMHRS likely contributed to a high proportion of people experiencing homelessness with severe mental illness. Given the differences in distribution of other variables, such as region of residence, we could not find a match for all people experiencing homelessness. This reflects major differences between homeless and non-homeless populations and reinforces the need to adjust for or match on important demographic and clinical characteristics.

### Implications

The findings of this work have implications for diabetes care and resource investment for people with diabetes experiencing homelessness. Healthcare providers are uniquely positioned to identify homelessness and work with their patients to reduce individual barriers to diabetes care; however, it is also important for organizations to promote low-barrier access to services and facilitate patient connections within the larger care system (32). Qualitative studies provide important context for the unmet care needs of people with diabetes experiencing homelessness. Population-level estimates can further quantify the need for tailored diabetes interventions and system changes to improve diabetes care for people experiencing homelessness.

Housing interventions have shown promise to mitigate the development of diabetes and related complications among people experiencing homelessness. One study reported housing vouchers were associated with reductions in the prevalence of obesity and diabetes among women and children living in high-poverty areas in urban centres (33). Another study linked supportive housing placements with reductions in diabetes diagnoses and improvements in diabetes care for adults experiencing homelessness (34). Access to housing remains a priority and should be supplemented with strategies to improve access to care and prevent complications for people with chronic diseases experiencing homelessness. Previous work highlights the importance of diabetes outreach programs in the community and shelter-based specialist care (35).

## Conclusion

Diabetes management is challenging for people experiencing homelessness, and poorer health outcomes are commonly reported. Yet the true increased risk in adverse health outcomes for this population has not been robustly described. This study provides population-level comparisons of the demographic and clinical characteristics of people with diabetes who use hospital services by housing status. This lays the foundation for future studies that seek to examine the impact of homelessness on diabetes outcomes at the population-level. This information should enable investment into diabetes care and program delivery for people experiencing homelessness, a group with unique needs and potentially avoidable excess risks of adverse diabetes outcomes.

**Table 1:** Demographic and clinical characteristics for people with diabetes who use hospital services in Ontario, comparing homeless and non-homeless groups prior to matching.

| Unmatched cohort of patients with diabetes | People with a history of homelessness (n=6944) | Non-homeless controls (n=1,069,493) | Standardized difference | P-value |
| --- | --- | --- | --- | --- |
| **Index dates** |  |  |  |  |
| 2006 | 502 (7.2%) | 77,253 (7.2%) | 0 | 0.583 |
| 2007 | 442 (6.4%) | 66,837 (6.2%) | 0 |  |
| 2008 | 416 (6.0%) | 61,642 (5.8%) | 0.01 |  |
| 2009 | 401 (5.8%) | 59,527 (5.6%) | 0.01 |  |
| 2010 | 375 (5.4%) | 56,194 (5.3%) | 0.01 |  |
| 2011 | 387 (5.6%) | 58,849 (5.5%) | 0 |  |
| 2012 | 411 (5.9%) | 62,539 (5.8%) | 0 |  |
| 2013 | 462 (6.7%) | 67,430 (6.3%) | 0.01 |  |
| 2014 | 469 (6.8%) | 69,079 (6.5%) | 0.01 |  |
| 2015 | 464 (6.7%) | 69,851 (6.5%) | 0.01 |  |
| 2016 | 530 (7.6%) | 81,733 (7.6%) | 0 |  |
| 2017 | 648 (9.3%) | 103,276 (9.7%) | 0.01 |  |
| 2018 | 1,137 (16.4%) | 183,467 (17.2%) | 0.02 |  |
| 2019 | 300 (4.3%) | 51,816 (4.8%) | 0.03 |  |
| **Demographic characteristics** |  |  |  |  |
| **Age** |  |  |  |  |
| Mean ± SD | 54.00 ± 14.24 | 65.63 ± 14.76 | 0.80 | <0.001 |
| 18-44 | 1,812 (26.1%) | 95,683 (8.9%) | 0.46 | <0.001 |
| 45-64 | 3,560 (51.3%) | 387,091 (36.2%) | 0.31 |  |
| $\geq 65$ | 1,572 (22.6%) | 586,719 (54.9%) | 0.70 | |
| **Sex** |  |  |  |  |
| Female | 2,348 (33.8%) | 521,303 (48.7%) | 0.31 | <0.001 |
| Male | 4,596 (66.2%) | 548,190 (51.3%) | 0.31 |  |
| **Geographical location (LHIN\*)** |  |  |  |  |
| Toronto Central | 1,761 (25.4%) | 76,203 (7.1%) | 0.51 | <0.001 |
| Central East | 703 (10.1%) | 136,220 (12.7%) | 0.08 |  |
| Hamilton Niagara Haldimand Brant | 666 (9.6%) | 118,711 (11.1%) | 0.05 |  |
| Southwest | 617 (8.9%) | 83,366 (7.8%) | 0.04 |  |
| Champlain | 542 (7.8%) | 95,609 (8.9%) | 0.04 |  |
| Central | 441 (6.4%) | 125,855 (11.8%) | 0.19 |  |
| Other | 2214 (31.9%) | 433,529 (40.5%) | - |  |
| **Clinical characteristics** |  |  |  |  |
| **Hospital encounter type**<br>(Index date +/-6 months) |  |  |  |  |
| Psychiatric inpatient admission to designated mental health bed (OMRHS*) | 2,559 (36.9%) | 14,487 (1.4%) | 1.01 | <0.001 |
| Other psychiatric and non-psychiatric inpatient admissions (DAD*) | 1,539 (22.2%) | 239,016 (22.3%) | 0.00 | 0.711 |
| Emergency department encounter (NACRS*) | 6,003 (86.4%) | 977,350 (91.4%) | 0.16 | <0.001 |
| **Mental illness** |  |  |  |  |
| Any mental illness | 3,397 (48.9%) | 26,032 (2.4%) | 1.26 | <0.001 |
| Psychotic disorder | 1,659 (23.9%) | 13,331 (1.2%) | 0.73 | <0.001 |
| Bipolar disorder | 867 (12.5%) | 13,718 (1.3%) | 0.45 | <0.001 |
| Other disorder | 3,256 (46.9%) | 148,584 (13.9%) | 0.77 | <0.001 |
| **Substance use disorders** |  |  |  |  |
| Alcohol use disorders | 1,021 (14.7%) | 6,724 (0.6%) | 0.55 | <0.001 |
| Non-alcohol substance use disorders | 1,189 (17.1%) | 7,328 (0.7%) | 0.60 | <0.001 |
| **Duration of diabetes** |  |  |  |  |
| <5 years | 1,416 (20.4%) | 155,688 (14.6%) | 0.15 | <0.001 |
| 5-10 years | 1,385 (19.9%) | 206,028 (19.3%) | 0.02 |  |
| 10-20 years | 1,645 (23.7%) | 268,913 (25.1%) | 0.03 |  |
| >20 years | 2,498 (36.0%) | 438,864 (41.0%) | 0.10 |  |
| **Type of diabetes** |  |  |  |  |
| Type I diabetes | 252 (3.6%) | 17,925 (1.7%) | 0.12 | <0.001 |
| Type II diabetes | 6,692 (96.4%) | 1,051,568 (98.3%) |  |  |
| **Major adverse cardiovascular event (MACE)** |  |  |  |  |
| Any MACE | 1,866 (26.9%) | 317,236 (29.7%) | 0.06 | <0.001 |
| Acute myocardial infarction | 840 (12.1%) | 158,559 (14.8%) | 0.08 | <0.001 |
| Congestive heart failure | 1,028 (14.8%) | 174,416 (16.3%) | 0.04 | <0.001 |
| Stroke | 688 (9.9%) | 107,187 (10.0%) | 0.00 | 0.752 |
| **Other chronic conditions** |  |  |  |  |
| Chronic Obstructive Pulmonary Disease | 1,062 (15.3%) | 99,380 (9.3%) | 0.18 | <0.001 |
| Connective tissue / rheumatic disease | 71 (1.0%) | 11,947 (1.1%) | 0.01 | 0.455 |
| Dementia | 203 (2.9%) | 31,152 (2.9%) | 0 | 0.958 |
| Hemiplegia or Paraplegia | 165 (2.4%) | 18,478 (1.7%) | 0.05 | <0.001 |
| HIV/AIDS | 47 (0.7%) | 674 (0.1%) | 0.10 | <0.001 |
| Mild Liver Disease | 547 (7.9%) | 17,773 (1.7%) | 0.29 | <0.001 |
| Moderate or Severe Liver Disease | 181 (2.6%) | 10,759 (1.0%) | 0.12 | <0.001 |
| Peptic Ulcer Disease | 215 (3.1%) | 26,736 (2.5%) | 0.04 | 0.002 |
| Primary Cancer | 155 (2.2%) | 43,101 (4.0%) | 0.10 | <0.001 |
| Metastatic Cancer | 88 (1.3%) | 31,276 (2.9%) | 0.12 | <0.001 |
| Renal Disease | 355 (5.1%) | 58,301 (5.5%) | 0.02 | 0.215 |

\* LHIN = Local Health Integration Network; OMHRS = Ontario Mental Health Reporting System; DAD = Discharge Abstract Database; NACRS: National Ambulatory Care Reporting System

**Table 2:** Baseline demographic and clinic characteristics for the matched cohorts, comparing demographic and clinical characteristics of patients experiencing homelessness to non-homeless and low-income controls.

| Matched cohort of patients with diabetes | People with a history of homelessness vs non-homeless controls |  |  |  | People with a history of homelessness vs low-income controls |  |  |  |
| --- | --- | --- | --- | --- | --- | --- | --- | --- |
|  | Homeless (n=5219) | Non-homeless controls (n=5219) | Standardized difference | Variance ratio | Homeless (n=4670) | Low-income controls (n=4670) | Standardized difference | Variance ratio |
| **Demographic characteristics** |  |  |  |  |  |  |  |  |
| **Age at index** |  |  |  |  |  |  |  |  |
| Mean ± SD | 55.20 ± 13.58 | 55.57 ± 13.69 | 0.027 | 0.984 | 55.76 ± 13.46 | 56.05 ± 13.58 | 0.021 | 0.983 |
| 18-44 | 1142 (21.9%) | 1142 (21.9%) | 0 | - | 941 (20.1%) | 941 (20.1%) | 0 | - |
| 45-64 | 2837 (54.4%) | 2837 (54.4%) |  |  | 2555 (54.7%) | 2555 (54.7%) |  |  |
| $\geq 65$ | 1240 (23.8%) | 1240 (23.8%) | | | 1174 (25.1%) | 1174 (25.1%) | | |
| **Sex** |  |  |  |  |  |  |  |  |
| Female | 1,688 (32.3%) | 1,737 (33.3%) | 0.02 | - | 1,511 (32.4%) | 1,572 (33.7%) | 0.028 | - |
| Male | 3,531 (67.7%) | 3,482 (66.7%) |  |  | 3,159 (67.6%) | 3,098 (66.3%) |  |  |
| **Geographical location (LHIN) \*** |  |  |  |  |  |  |  |  |
| Toronto Central | 1,420 (27.2%) | 1,420 (27.2%) | 0 | - | 1,305 (27.9%) | 1,305 (27.9%) | 0 | - |
| Central East | 547 (10.5%) | 547 (10.5%) |  |  | 495 (10.6%) | 495 (10.6%) |  |  |
| Hamilton Niagara Haldimand Brant | 514 (9.8%) | 514 (9.8%) |  |  | 461 (9.9%) | 461 (9.9%) |  |  |
| Southwest | 448 (8.6%) | 448 (8.6%) |  |  | 414 (8.9%) | 414 (8.9%) |  |  |
| Champlain | 430 (8.2%) | 430 (8.2%) |  |  | 389 (8.3%) | 389 (8.3%) |  |  |
| Central | 334 (6.4%) | 334 (6.4%) |  |  | 281 (6.0%) | 281 (6.0%) |  |  |
| Other | 1526 (29.2%) | 1526 (29.2%) |  |  | 1325 (28.4%) | 1325 (28.4%) |  |  |
| **Clinical characteristics** |  |  |  |  |  |  |  |  |
| **Hospital encounter type**<br>(Index date +/-6 months) |  |  |  |  |  |  |  |  |
| Psychiatric inpatient admission to designated mental health bed (OMRHS*) | 1,254 (24.0%) | 1,254 (24.0%) | 0 | - | 882 (18.9%) | 882 (18.9%) | 0 | - |
| Other psychiatric and non-psychiatric inpatient admissions (DAD*) | 1,149 (22.0%) | 1,149 (22.0%) | 0 | - | 992 (21.2%) | 992 (21.2%) | 0 | - |
| Emergency department encounter (NACRS*) | 4,911 (94.1%) | 4,911 (94.1%) | 0 | - | 4,428 (94.8%) | 4,428 (94.8%) | 0 | - |
| **Chronic conditions** |  |  |  |  |  |  |  |  |
| Any mental illness | 3,900 (74.7%) | 3,900 (74.7%) | 0 | - | 3,388 (72.5%) | 3,388 (72.5%) | 0 | - |
| Acute myocardial infarction | 604 (11.6%) | 646 (12.4%) | 0.025 | - | 574 (12.3%) | 512 (11.0%) | 0.041 | - |
| Congestive heart failure | 699 (13.4%) | 838 (16.1%) | 0.075 | - | 748 (16.0%) | 593 (12.7%) | 0.095 | - |
| Stroke | 426 (8.2%) | 552 (10.6%) | 0.083 | - | 502 (10.7%) | 354 (7.6%) | 0.110 | - |
| **Duration of diabetes** |  |  |  |  |  |  |  |  |
| <5 years | 1051 (20.1%) | 1029 (19.7%) | 0.011 | - | 812 (17.4%) | 851 (18.2%) | 0.022 | - |
| 5-10 years | 1,040 (19.9%) | 1,043 (20.0%) | 0.001 | - | 935 (20.0%) | 888 (19.0%) | 0.025 | - |
| 10-20 years | 1,209 (23.2%) | 1,213 (23.2%) | 0.002 | - | 1,151 (24.6%) | 1,175 (25.2%) | 0.012 | - |
| >20 years | 1,919 (36.8%) | 1,934 (37.1%) | 0.006 | - | 1,772 (37.9%) | 1,756 (37.6%) | 0.007 | - |
| **Type of diabetes** |  |  |  |  |  |  |  |  |
| Type I diabetes | 127 (2.4%) | 109 (2.1%) | 0.023 | - | 80 (1.7%) | 90 (1.9%) | 0.016 | - |
| Type II diabetes | 5092 (97.6%) | 5110 (97.9%) |  |  | 4590 (98.3%) | 4580 (98.1%) |  |  |

\* LHIN = Local Health Integration Network; OMRHS = Ontario Mental Health Reporting System; DAD = Discharge Abstract Database; NACRS: National Ambulatory Care Reporting System

## Data Availability

The datasets generated and analyzed during the current study are not publicly available due to data sharing agreements and privacy policies that prohibit ICES from sharing the dataset publicly. The datasets supporting the conclusions of this article are held securely in coded form at ICES. Upon reasonable request, confidential access may be granted to those who meet pre-specified criteria.

## Ethics Statement

ICES is a prescribed entity under Ontario’s Personal Health Information Protection Act (PHIPA). Section 45 of PHIPA authorizes ICES to collect personal health information, without consent, for the purpose of analysis or compiling statistical information with respect to the management of, evaluation or monitoring of the allocation of resources to or planning for all or part of the health system. Projects that use data collected by ICES under section 45 of PHIPA, and use no other data, are exempt from REB review. The use of the data in this project is authorized under section 45 and approved by ICES’ Privacy and Legal Office. This project has also been approved by the Research Ethics Board at the University of Calgary.

