## Supplemental Table S1 for "Characteristics of People with Type I or Type II Diabetes with and without a History of Homelessness: A Population-based Cohort Study"

Supplemental Table S1: Description of administrative healthcare databases.

| Databases ^a^ | Description | Important variables |
| --- | --- | --- |
| RPDB: Registered persons database | A registry of all Ontario residents who have ever received a provincial health card number | Date of birth, sex, geographical region, neighborhood income quintile, dates of eligibility for health coverage, date of death |
| ODD: Ontario Diabetes Dataset ^b^ | A registry of physician-diagnosed diabetes mellitus, based on a validated algorithm. The ODD inclusion criteria are two outpatient physician claims for diabetes within two years or one hospital admission for diabetes. | Duration of diabetes (date of entry into the ODD) |
| DAD: Discharge Abstract Database ^b^ | Source of data on separations from acute care inpatient institutions including day surgery, chronic, rehabilitation and psychiatric institutions | Homelessness, inpatient admissions, major adverse cardiovascular events, revascularization procedures |
| OMHRS: Ontario Mental Health Reporting System ^c^ | Source of data for inpatient admissions to designated adult mental health beds in Ontario | Homelessness, psychiatric inpatient admissions, mental illness, substance use disorders |
| NACRS: National Ambulatory Care Reporting System ^d^ | Source of data on emergency and ambulatory care services in the hospital and community | Homelessness, emergency department visits |

a. ICES Data Dictionary: <https://datadictionary.ices.on.ca/Applications/DataDictionary/Default.aspx>
b. Data Quality Documentation, Discharge Abstract Database: <https://www.cihi.ca/sites/default/files/document/dad-data-quality-current-year-information-2020-2021-en.pdf>
c. Data Quality Documentation, Ontario Mental Health Reporting System: <https://www.cihi.ca/sites/default/files/document/omhrs-data-quality-2020-2021-report-en.pdf>
d. Data Quality Documentation, National Ambulatory Care Reporting System: <https://www.cihi.ca/sites/default/files/document/nacrs-data-quality-current-year-information-2020-2021-en.pdf>
