## Supplemental Table S2 for "Characteristics of People with Type I or Type II Diabetes with and without a History of Homelessness: A Population-based Cohort Study"

Supplemental Table S2: List of administrative data elements to identify homelessness in hospital databases ^a^

| *Database* | *Indicator Code* | *Coding Description* |
| --- | --- | --- |
| *Ontario Mental Health Reporting System (OMHRS)* | *PRIOR_RESIDENCE = 6* | *Homeless (with or without shelter)* |
|  | *USUAL_RESIDENCE = 8* | *Homeless (with or without shelter)* |
|  | *ADMITFROM = 8* | *Homeless (with or without shelter)* |
|  | *DISCHLIVING = 8* | *Homeless (with or without shelter)* |
|  | *P5_Retired_2009 = 6* | *Homeless (with or without shelter)* |
|  | *PSTLCODE = XX* | *No fixed address* |
|  | *PREDX10CODE1-11 = Z590 or Z591* | *Homelessness or inadequate housing* |
|  | *POSTDX10CODE1-24 = Z590 or Z591* | *Homelessness or inadequate housing* |
| *Discharge Abstract Database (DAD)* | *HOMELESS = Y* | *Yes, homeless* |
|  | *INSTTYPE = SH* | *Supportive housing* |
|  | *PSTLCODE = XX* | *No fixed address* |
|  | *DX10CODE1-25 = Z590 or Z591* | *Homelessness or inadequate housing* |
|  | *CMGDIAG = Z590 or Z591* | *Homelessness or inadequate housing* |
| *National Ambulatory Care Reporting System (NACRS)* | *RESTYPE = 3 or 4* | *Homeless or shelter* |
|  | *PSTLCODE = XX* | *No fixed address* |
|  | *DX10CODE1-25 = Z590 or Z591* | *Homelessness or inadequate housing* |

a. Modified from: Richard L, Hwang SW, Forchuk C, Nisenbaum R, Clemens K, Wiens K, et al. Validation study of health administrative data algorithms to identify individuals experiencing homelessness and estimate population prevalence of homelessness in Ontario, Canada. BMJ Open. 2019;9(10):e030221.
