## Supplemental Figure S1 for "Characteristics of People with Type I or Type II Diabetes with and without a History of Homelessness: A Population-based Cohort Study"

Supplemental Figure S1: A summary of the methods to create a matched cohort of people with diabetes with a history of homelessness and non-homeless controls.

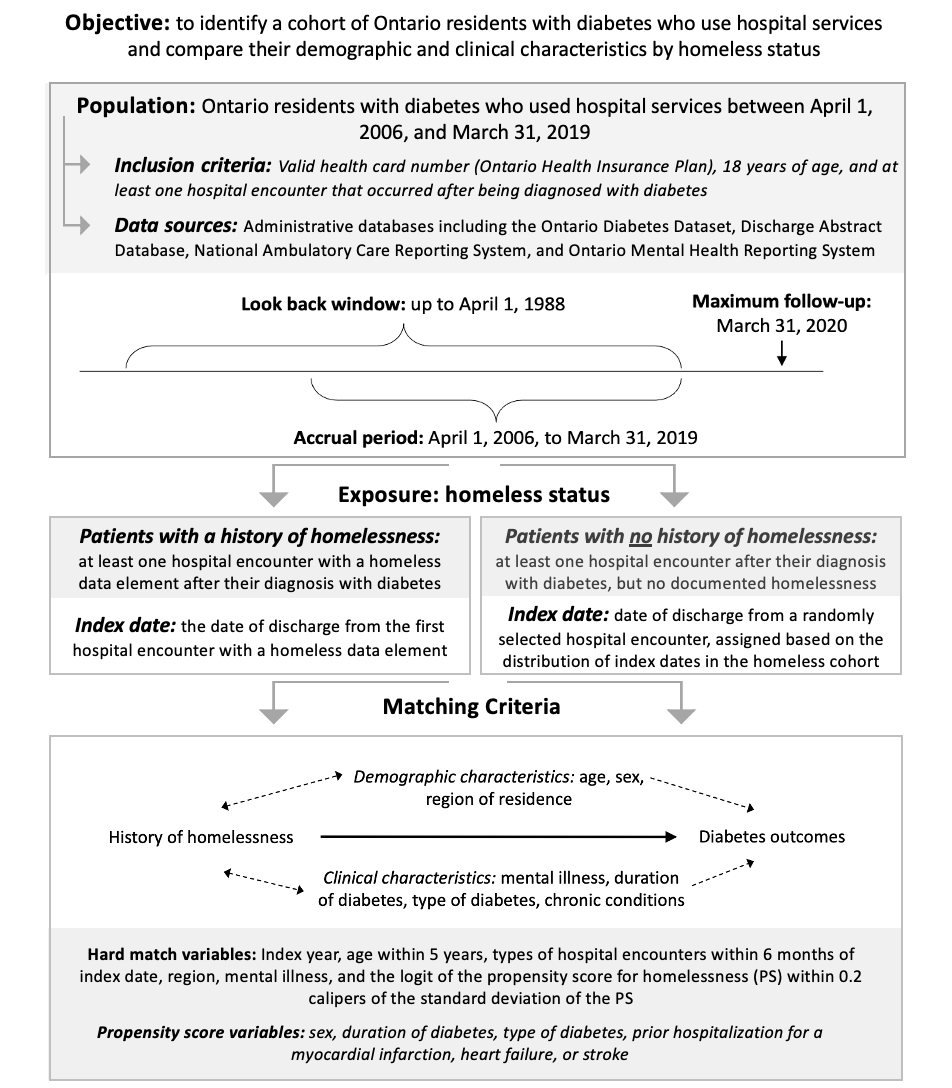
