## Supplemental Table S3 for "Characteristics of People with Type I or Type II Diabetes with and without a History of Homelessness: A Population-based Cohort Study"

Supplemental Table S3: Comparing the demographic and clinical characteristics for the included and excluded samples of people with a history of homelessness (matched versus not matched).

| **Unmatched cohort of patients with diabetes** | **Matched (n=5219)** | **Not matched (n=1725)** | **Standardized difference** | **P-value**  **(a=0.05)** |
| --- | --- | --- | --- | --- |
| **Index dates** |  |  |  |  |
| 2006 | 401 (7.7%) | 101 (5.9%) | 0.07 | <.001 |
| 2007 | 331 (6.3%) | 111 (6.4%) | 0 |  |
| 2008 | 283 (5.4%) | 133 (7.7%) | 0.09 |  |
| 2009 | 274 (5.3%) | 127 (7.4%) | 0.09 |  |
| 2010 | 233 (4.5%) | 142 (8.2%) | 0.15 |  |
| 2011 | 253 (4.8%) | 134 (7.8%) | 0.12 |  |
| 2012 | 286 (5.5%) | 125 (7.2%) | 0.07 |  |
| 2013 | 329 (6.3%) | 133 (7.7%) | 0.06 |  |
| 2014 | 344 (6.6%) | 125 (7.2%) | 0.03 |  |
| 2015 | 356 (6.8%) | 108 (6.3%) | 0.02 |  |
| 2016 | 399 (7.6%) | 131 (7.6%) | 0 |  |
| 2017 | 513 (9.8%) | 135 (7.8%) | 0.07 |  |
| 2018 | 988 (18.9%) | 149 (8.6%) | 0.3 |  |
| 2019 | 229 (4.4%) | 71 (4.1%) | 0.01 |  |
| **Demographic characteristics** |  |  |  |  |
| **Age** |  |  |  |  |
| Mean ± SD | 55.20 ± 13.58 | 50.39 ± 15.53 | 0.33 | <0.001 |
| Median (IQR) | 55 (46-64) | 50 (39-61) | 0.33 | <0.001 |
| **Sex** |  |  |  |  |
| Female | 1,688 (32.3%) | 660 (38.3%) | 0.12 | <0.001 |
| Male | 3,531 (67.7%) | 1,065 (61.7%) | 0.12 |  |
| **Geographical Location (LHIN*)** |  |  |  |  |
| Toronto Central | 1,420 (27.2%) | 341 (19.8%) | 0.18 | <0.001 |
| Central East | 547 (10.5%) | 156 (9.0%) | 0.05 |  |
| Hamilton Niagara Haldimand Brant | 514 (9.8%) | 152 (8.8%) | 0.04 |  |
| Southwest | 448 (8.6%) | 169 (9.8%) | 0.04 |  |
| Champlain | 430 (8.2%) | 112 (6.5%) | 0.07 |  |
| Central | 334 (6.4%) | 107 (6.2%) | 0.01 |  |
| Other | 1526 (29.2%) | 688 (39.9%) | - |  |
| **Clinical characteristics** |  |  |  |  |
| **Hospital use**  (Index date +/-6 months) |  |  |  |  |
| Psychiatric inpatient admission to designated mental health bed (OMRHS*) | 1,254 (24.0%) | 1,305 (75.7%) | 1.21 | <0.001 |
| Other psychiatric and non-psychiatric inpatient admissions (DAD*) | 1,149 (22.0%) | 390 (22.6%) | 0.01 | 0.607 |
| Emergency department encounter (NACRS*) | 4,911 (94.1%) | 1,092 (63.3%) | 0.81 | <0.001 |
| **Mental illness** |  |  |  |  |
| Psychotic disorder | 987 (18.9%) | 4,232 (81.1%) | 0.45 | <0.001 |
| Bipolar disorder | 500 (9.6%) | 367 (21.3%) | 0.33 | <0.001 |
| Other disorder | 2,261 (43.3%) | 995 (57.7%) | 0.29 | <0.001 |
| **Substance use disorders** |  |  |  |  |
| Alcohol use disorders | 817 (15.7%) | 204 (11.8%) | 0.11 | <0.001 |
| Non-alcohol use disorders | 891 (17.1%) | 298 (17.3%) | 0.01 | 0.846 |
| **Duration of diabetes** |  |  |  |  |
| <5 years | 1,029 (19.7%) | 387 (22.4%) | 0.07 | 0.004 |
| 5-10 years | 1,043 (20.0%) | 342 (19.8%) | 0.00 |  |
| 10-20 years | 1,213 (23.2%) | 432 (25.0%) | 0.04 |  |
| >20 years | 1,934 (37.1%) | 564 (32.7%) | 0.09 |  |
| **Type of diabetes** |  |  |  |  |
| Type I diabetes | 109 (2.1%) | 143 (8.3%) | 0.28 | <0.001 |
| Type II diabetes | 5,110 (97.9%) | 1,582 (91.7%) |  |  |
| **Major adverse cardiovascular event (MACE)** |  |  |  |  |
| Any MACE | 1,482 (28.4%) | 384 (22.3%) | 0.14 | <0.001 |
| Acute myocardial infarction | 646 (12.4%) | 194 (11.2%) | 0.04 | 0.212 |
| Congestive heart failure | 838 (16.1%) | 190 (11.0%) | 0.15 | <0.001 |
| Stroke | 552 (10.6%) | 136 (7.9%) | 0.09 | 0.001 |
| **Other Chronic Conditions** |  |  |  |  |
| Chronic Obstructive Pulmonary Disease | 808 (15.5%) | 254 (14.7%) | 0.02 | 0.449 |
| Connective tissue / Rheumatic disease | 59 (1.1%) | 12 (0.7%) | 0.05 | 0.120 |
| Dementia | 149 (2.9%) | 54 (3.1%) | 0.02 | 0.556 |
| Hemiplegia or Paraplegia | 137 (2.6%) | 28 (1.6%) | 0.07 | 0.018 |
| HIV/AIDS | 35 (0.7%) | 12 (0.7%) | 0 | 0.912 |
| Mild Liver Disease | 433 (8.3%) | 114 (6.6%) | 0.06 | 0.024 |
| Moderate or Severe Liver Disease | 164 (3.1%) | 17 (1.0%) | 0.15 | <0.001 |
| Peptic Ulcer Disease | 158 (3.0%) | 57 (3.3%) | 0.02 | 0.565 |
| Primary Cancer | 112 (2.1%) | 43 (2.5%) | 0.02 | 0.298 |
| Metastatic Cancer | 69 (1.3%) | 19 (1.1%) | 0.02 | 0.478 |
| Renal Disease | 287 (5.5%) | 68 (3.9%) | 0.07 | 0.011 |

* LHIN = Local Health Integration Network; OMHRS = Ontario Mental Health Reporting System; DAD = Discharge Abstract Database; NACRS: National Ambulatory Care Reporting System
